# Type 1 interferon auto-antibodies are elevated in patients with decompensated liver cirrhosis

**DOI:** 10.1101/2022.12.14.22283445

**Authors:** Gordon Greville, Sinead Cremen, Shauna O’Neill, Sarah Azarian, Gareth Brady, William McCormack, Olivier Touzelet, David Courtney, Ultan Power, Paul Dowling, Tom K Gallagher, Connor GG Bamford, Mark W Robinson

## Abstract

Understanding the biological basis of clinical risk factors for severe COVID-19 is required to ensure at-risk patient populations receive appropriate clinical care. Patients with decompensated liver cirrhosis, in particular those classified as Childs-Pugh class B and C, are at increased risk of severe COVID-19 upon infection with SARS-CoV-2. The biological mechanisms underlying this are unknown. We hypothesised this may be due to changes in expression levels of intrinsic antiviral proteins within the serum as well as alterations in the innate immune response to SARS-CoV-2 infection. We identified significant alterations in the serum proteome of patients with more severe liver disease and an increased frequency of auto-antibodies capable of neutralising type I interferons. No difference in SARS-CoV-2 pseudoparticle infection or live SARS-CoV-2 virus infection was observed with serum from decompensated cirrhotic patients. Principal component analysis of the serum proteome identified two main clinical parameters associated with serum proteome changes – aetiology and MELD-Na score. Among patients with MELD-Na scores >20 we detected significant inhibition of IFN-α2b and IFN-α8 signalling but not IFN-β1a, mediated by auto-antibodies. Our results suggest pre-existing neutralising auto-antibodies targeting type I IFN may increase the likelihood of severe COVID-19 in chronic liver disease patients upon SARS-CoV-2 infection and may also be of relevance to other viral infections in this patient population.

## Introduction

A number of risk factors associated with the progression to severe COVID-19 upon infection with severe acute respiratory coronavirus 2 (SARS-CoV-2) have been described. These include factors such as age, sex, diabetes, genetics, cardiac disease, class III obesity, and cancer.[1, 2]. Pre-existing chronic liver disease (CLD) is also an important risk factor for mortality during severe COVID-19 [1, 3, 4]. Early retrospective studies of patients with liver cirrhosis and SARS-CoV-2 infection identified high rates of mortality [5, 6]. Subsequent data from international registries of COVID-19 in chronic liver disease patients identified increased mortality in patients with high Child-Pugh and MELD scores [7, 8]. Deaths occurred in 8% of SARS-CoV-2 infected non-cirrhotic patients but 32% of infected cirrhotic patients, and this increased to 51% in Child-Pugh class C patients [8].

The biological mechanisms underlying these various risk factors are complex and remain only partially resolved. One described biological mechanism linked to aging is the presence of auto-antibodies capable of neutralising type I interferons [9, 10]. The presence of type interferon (IFN) auto-antibodies (AAb) increases with age – in individuals over the age of 75 between 2-8% possess auto-antibodies capable of neutralising 100 pg/mL of IFN-α2 in human embryonic kidney (HEK)293T type I IFN reporter assays [9]. Neutralising AAb against type I IFNs are enriched in individuals with severe COVID-19, being present in 18% of individuals who die from SARS-CoV-2 infection [9], and are also present in approximately 5% of life-threatening influenza pneumonia cases in individuals younger than 70 years old [11].

In the context of pre-existing chronic liver disease a number of mechanisms have been postulated as possible causes of the increased risk for severe COVID-19 [3]. These include hypercoagulation driven by systemic inflammation, alterations in the gut-liver axis, acute hepatic decompensation due to infection of hepatocytes, altered innate immunity enhancing viral replication, and reduced adaptive immune responses due to cirrhosis-associated immune dysfunction. We hypothesised that cirrhosis-associated immune dysfunction may influence SARS-CoV-2 infection in two ways: firstly, by altering innate antiviral proteins in the serum, which can inhibit SARS-CoV-2, thereby enhancing viral infectivity; and secondly, by dysregulating inflammatory signalling pathways resulting in enhanced pro-inflammatory signals. In the present study we demonstrate that viral infectivity is not altered in the presence of serum from patients with decompensated liver cirrhosis, but these patients do have elevated levels of auto-antibodies capable of neutralising type I interferons, supporting our second hypothesis.

## Methods

### Participant recruitment and processing

Study participants were approached after being referred for transplant assessment at the Irish National Liver Transplant Centre at St Vincent’s Hospital from December 2019 to December 2020. This was prior to the widespread circulation of SARS-CoV-2 within Ireland and the development of SARS-CoV-2 vaccines. Patients eligible for inclusion were those over 18 years who were undergoing liver transplant assessment regardless of disease aetiology. Patients were reviewed either at outpatient clinic appointments or as inpatients. Prior to taking part in the study, they were fully informed on the procedures and gave written consent. Blood samples were taken at the time of clinical assessment for clinical laboratory analysis and research. Blood for serum isolation was collected in a serum blood collection tube with no anticoagulant or preservative. Blood was allowed to clot and then centrifuged at 1000 rcf for 10 minutes. Serum was collected and centrifuged a second time before being stored in aliquots at -70 C.

### SARS-CoV-2 serological screening

All participants were screened for the presence of serological reactivity against SARS-CoV-2 proteins in their serum. Serum IgG and IgM responses against the spike protein of SARS-CoV-2 was performed as previously described [12]. Cut-off values were determined with a reference positive control sample. Serum IgG responses against the nucleocapsid protein of SARS-CoV-2 was performed as previously described [13]. Absorption at 450 nm was measured using a CLARIOstar® Plus plate reader with a reference positive control sample utilised as cut-off. Seroreactivity was subsequently confirmed in a SARS-CoV-2 pseudoparticle assay as detailed below.

### Pseudoparticle assays

The effect of serum on SARS-CoV-2 entry was assessed using a lentivirus-based pseudotype system, using the reference sequence of Wuhan-Hu-1 spike, as described elsewhere [14]. An expression vector was constructed in pcDNA3.1(-) to produce spike lacking the C-terminal 18 amino acids of the cytoplasmic tail [15], based off spike plasmids kindly provided by Prof. Nigel Temperton (Medway School of Pharmacy). To produce pseudoparticles, the spike plasmid was co-transfected into near confluent wild-type HEK-293T cells along with packaging (p8.91) and firefly luciferase reporter-expressing lentivirus genome (pCSFLW) plasmids at a ratio of 1:5:5. Plasmid were mixed with lipofectamine 2000 reagent as per the manufacturer’s instructions and incubated with cells overnight in OptiMEM before switching to standard growth media (high glucose DMEM with 10% FCS and 1% pen/strep). At 2 days post-transfection, conditioned medium containing pseudoparticles was harvested, clarified, and filtered using a 0.45 m filter. Pseudoparticles (25 l) were incubated with serum (50 l at a final dilution of 1:3 dilution) at 37 °C for 1 hr, before addition of target cells (50,000 per well of a 96-well plate, in an equal volume) to assess infectivity. Target cells used were lentiviral transduced HEK-293T cells expressing an EGFP fusion protein linked to human ACE2 (plasmids kindly provided by Dr Jacob Yount, Ohio State University) and human TMPRSS2 [16]. Firefly luciferase activity was measured using luciferase assay system detection kit (Promega), as per manufacturer’s instructions, and read using a Mithras LB940 (Berthold) plate reader 2 days post-transduction. Controls used included DMEM, foetal calf serum (FCS), and normal human serum. Data presented represent two independent experimental repeats.

### Live virus assay

For all SARS-CoV-2 work using infectious virus, experiments were carried out in a dedicated BSL3 facility at Queen’s University Belfast. The virus strain used was “BT20.1”, which is pre-variant of concern isolate carrying the D614G mutation in spike and retaining an intact polybasic cleavage site [17]. Modified human airway epithelial A549 cells expressing human ACE2 and TMPRSS2 [18] (kindly provided by Dr Suzannah Rihn, MRC-University of Glasgow Centre for Virus Research) were used in well clearance infection assays exploiting the cytopathic nature of this isolate. This assay measures cell lysis, so a higher clearance of the modified A549 monolayer indicates higher viral replication. Cell monolayers (∼90% confluent) were infected with virus at an MOI of 0.1 plaque-forming units per cell, and 2 h post infection serum was added at a final dilution of 1:3. At 2 days post-infection cells were fixed before staining with crystal violet. Stained wells were visualised using a CELIGO imaging cytometer (Nexelom) and images analysed in Fiji [19], using white pixel intensity quantification. Clearance was quantified as a percentage using an untreated control sample as the baseline for no clearance (0%) and an infected well treated with DMEM alone as the maximum clearance (100%). Data presented represent two independent experimental repeats.

### Mass spectrometry analysis

Serum samples were immunodepleted of albumin and IgG using Proteome Purify 2 Human Serum Protein Immunodepletion Resin (R&D). The removal of these high abundance serum proteins enhanced the proteomic detection of less abundant proteins present in the sample. After reduction with dithiothreitol and iodoacetic acid-mediated alkylation, digestion was performed using trypsin overnight at 37°C. Digested immunodepleted samples were loaded onto a Q-Exactive high-resolution accurate mass spectrometer connected to a Dionex Ultimate 3000 (RSLCnano) chromatography system (ThermoFisher Scientific, Hemel Hempstead, UK). Sample loading was conducted by an auto-sampler onto a C18 trap column (C18 PepMap, 300 μm id × 5 mm, 5 μm particle size, 100 Å pore size; ThermoFisher Scientific). The trap column was switched on-line with an analytical Biobasic C18 Picofrit column (C18 PepMap, 75 μm id × 50 cm, 2 μm particle size, 100 Å pore size: Dionex). Data were acquired with Xcalibur software (Thermo Fisher Scientific). Data analysis, processing and visualisation was performed using MaxQuant v1.5.2.8 (http://www.maxquant.org) and Perseus v.2.0.7.0 (www.maxquant.org/) software. The Andromeda search engine was used to explore the detected features against the UniProtKB/SwissProt database for *Homo sapiens*. The false discovery rate (FDR) was set to 1% for both peptides and proteins using a target-decoy approach. The intensities were log2 transformed, with proteins filtered based of detection in >85% of samples and data imputation was performed to replace missing values.

### Cytokine arrays

Expression of inflammatory cytokines were assessed using the LEGENDplex Human Inflammation Panel 1 (BioLegend), as per manufacturer’s instructions. Data was acquired on a BD Accuri C6 (BD Biosciences).

### Type I IFN reporter assays

Highly-sensitive commercially available human monocytic THP-1 dual reporter cells (InvivoGen), which express a secreted luciferase (Lucia) under the control of five IFN-stimulated response elements (ISREs), were cultured in RPMI 1640 (Gibco, ThermoFisher), 2 mM L-glutamine, 25 mM HEPES, 10% heat-inactivated fetal bovine serum, 100 μg/ml normocin, and penicillin-streptomycin at 37°C in 5% CO_2_. Selection pressure was maintained with 10 μg/ml of blasticidin and 100 μg/ml of zeocin added every other passage. Patient serum, at a final dilution of 1 in 40, was combined with either IFN-α2b (InvivoGen; 500 pg/mL), IFN-α8 (InvivoGen; 50 pg/mL) or IFN-β1a (InvivoGen; 100 pg/mL), and incubated with THP-1 dual reporter cells for 18 h at 37°C. These type I IFNs were selected as they show either intermediate (IFN-α2b) or high (IFN-α8) antiviral activity against SARS-CoV-2 and IFN-β1a is expressed by a wide range of cell types, compared to the restricted expression of IFN-α [20]. The concentrations of the type I IFNs were selected to be approximately half the IC_50_ in the THP-1 dual reporter cell system. Luminescence intensity was measured with a CLARIOstar® Plus microplate reader (BMG Labtech) and luciferase activity values were normalized against IFN-only positive and media-only negative controls and expressed as a percentage. Data presented represent two independent experimental repeats.

### IgG depletion

IgG was purified from serum using the NAb™ Protein G Spin kit (ThermoFisher Scientific), as per manufacturer’s instructions. The IgG-depleted flow-through was collected along with the purified IgG. The concentration of purified IgG was quantified using a NanoDrop™ 2000 Spectrophotometer (ThermoFisher Scientific). Purified IgG and IgG-depleted serum were assessed for IFN-α2b inhibition as described above. A final concentration of 200 μg/mL/well was selected for purified IgG, representative of the concentration of IgG in diluted serum. The IgG-depleted serum was analysed at a 1 in 40 dilution alongside the original serum sample.

### IFN inhibitory IgG ELISA

To assess the binding capacity of serum purified IgG to IFN-α2b, Nunc 96 well Maxisorp plates (ThermoFisher Scientific) were coated with IFN-α2b overnight at 4°C. Plates were blocked and incubated for 1 h at room temperature with 20 μg of serum purified IgG or a 1:40 dilution of healthy plasma. Plates were washed with 0.05% PBS Tween20 then incubated for 1 h with 500 ng/mL horse radish peroxidase-conjugated goat anti-human IgG (ThermoFisher Scientific). TMB (BioLegend) was added to each well and once colour developed the reaction was stopped by the addition of 1 M sulphuric acid. Absorbance at 450 nm was read on CLARIOstar® Plus microplate reader (BMG Labtech).

### Statistical analysis

Statistical analysis was performed using GraphPad Prism (version 9.3.0), using either parametric or non-parametric analysis as indicated, depending on the data distributions. Statistical analysis of mass spectrometry data was done using Perseus, utilising t-test to compare between two groups and a significant cut-off of q < 0.1. Principal component analysis of mass spectrometry data was performed using GraphPad Prism (version 9.3.0). Pathway analysis of mass spectrometry data was performed using ShinyGO v0.76 [21] using all detected proteins as a background list, a FDR cut-off of 0.05 and a maximum pathway size of 100.

## Results

### Identification of SARS-CoV-2 naïve individuals undergoing liver transplant assessment

A cohort of 56 individuals undergoing transplant assessment at the Irish National Liver Transplant Centre at St Vincent’s Hospital from December 2019 to December 2020 were assessed for serological responses against SARS-CoV-2 spike (S) and nucleocapsid (N) antigens (**Supplementary Figure 1**). S-antigen IgG or IgM reactivity was evident in 3 individuals, suggesting previous asymptomatic/unreported infection with SARS-CoV-2 (**Supplementary Figure 1A and B**). Two of these individuals also demonstrated N-antigen IgG reactivity (**Supplementary Figure 1C**), and all three demonstrated viral neutralisation using SARS-CoV-2 pseudoparticle assays (**Supplementary Figure 1D**). Upon exclusion of these three seropositive individuals, a total of 53 SARS-CoV-2 unexposed and unvaccinated individuals with varying degrees of chronic liver disease were included in the study as detailed in **Table 1**.

**Table 1:**
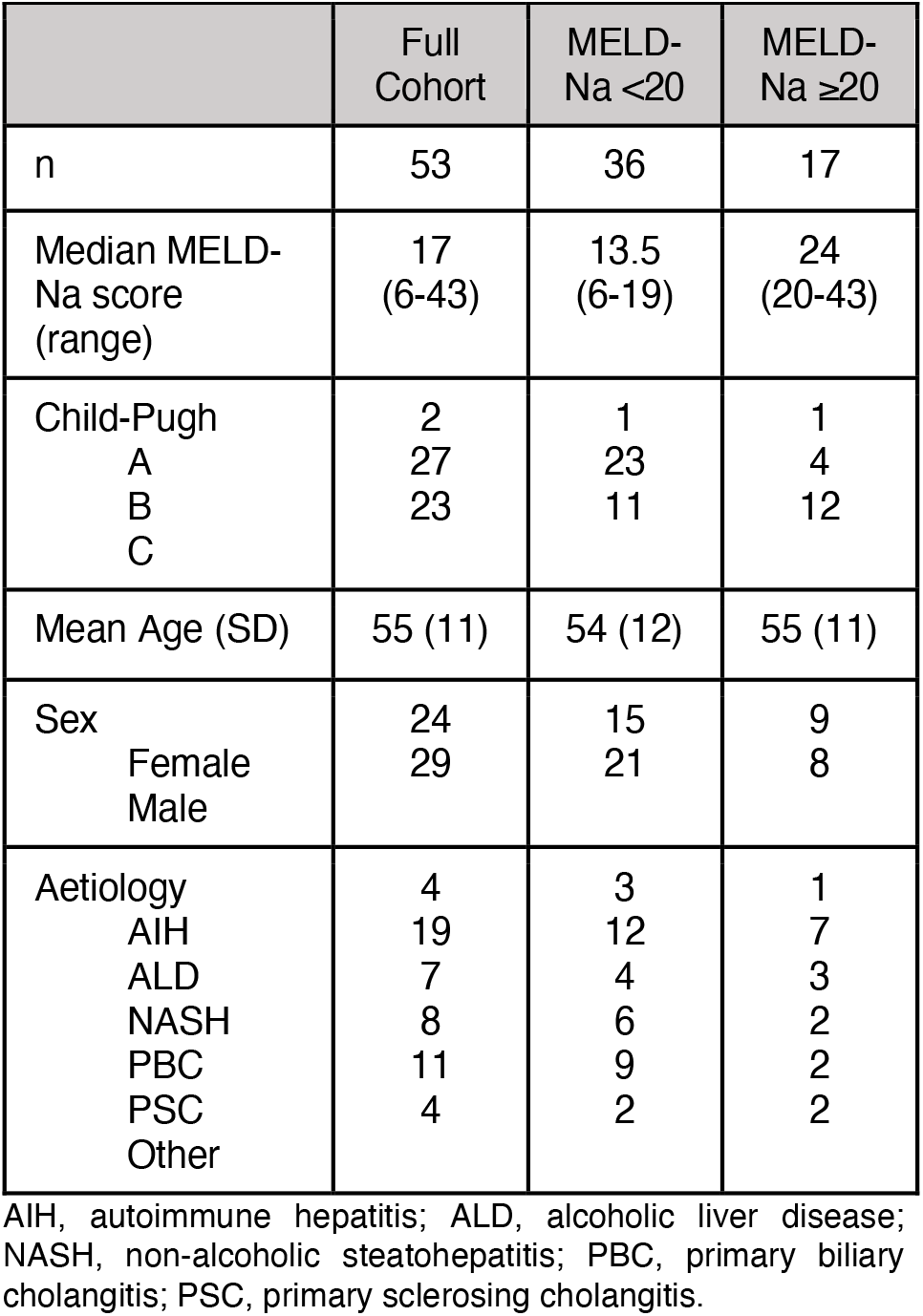
Clinical demographics of patient cohort.

These 53 individuals presented for liver transplant assessment with a range of underlying aetiologies, including alcoholic liver disease (ALD; n=19), non-alcoholic steatohepatitis (NASH; n=7), autoimmune hepatitis (AIH; n=4), primary biliary cholangitis (PBC; n=8), and primary sclerosing cholangitis (PSC; n=11). The total cohort included 45% females and had a mean age of 55 years, in line with national Irish trends for liver transplantation. To explore the biological mechanisms underlying the increased mortality upon SARS-CoV-2 infection in patients decompensated liver cirrhosis [7, 8], the cohort was divided on the basis of MELD-Na scores. Amongst the 53 individuals a total of 36 had MELD-Na scores less than 20 and within this group the majority of patients were classified as Child-Pugh class B (64%). A further 17 individuals had MELD-Na scores greater or equal to 20, amongst whom the majority were classified as Child-Pugh class C (71%). Within the MELD-Na ≥20 group there was a disease aetiology bias with a higher frequency of individuals with ALD and fewer individuals with an autoimmune diagnosis, in particular PSC.

### Serum from patients with decompensated liver disease does not alter viral replication or infectivity

Serum from patients within the cohort were screened using a SARS-CoV-2 pseudoparticle assay (**Figure 1A**). The addition of serum from different individuals resulted in wide variation in pseudoparticle infectivity, with serum from some individuals inhibiting infection while others enhanced infection. Pre-incubation of diluted serum from patients with MELD-Na ≥20 with SARS-CoV-2 pseudoparticles did not result in significantly different pseudoparticle infectivity compared to patients with MELD-Na <20 (**Figure 1B**). MELD-Na <20 and MELD-Na ≥20 groups had a median percentage pseudoparticle infectivity of 79.9% and 80.1% (p=0.371), respectively. There was no significant correlation between pseudoparticle infectivity and MELD-Na scores across the full cohort (**Figure 1C**). Similar to the results when individuals were grouped by MELD-Na scores, no significant difference in pseudoparticle infectivity was evident between patients classified as either Child-Pugh class B or C, nor between patients with different disease aetiologies (**Figure 1D, E**).

**Figure 1:**
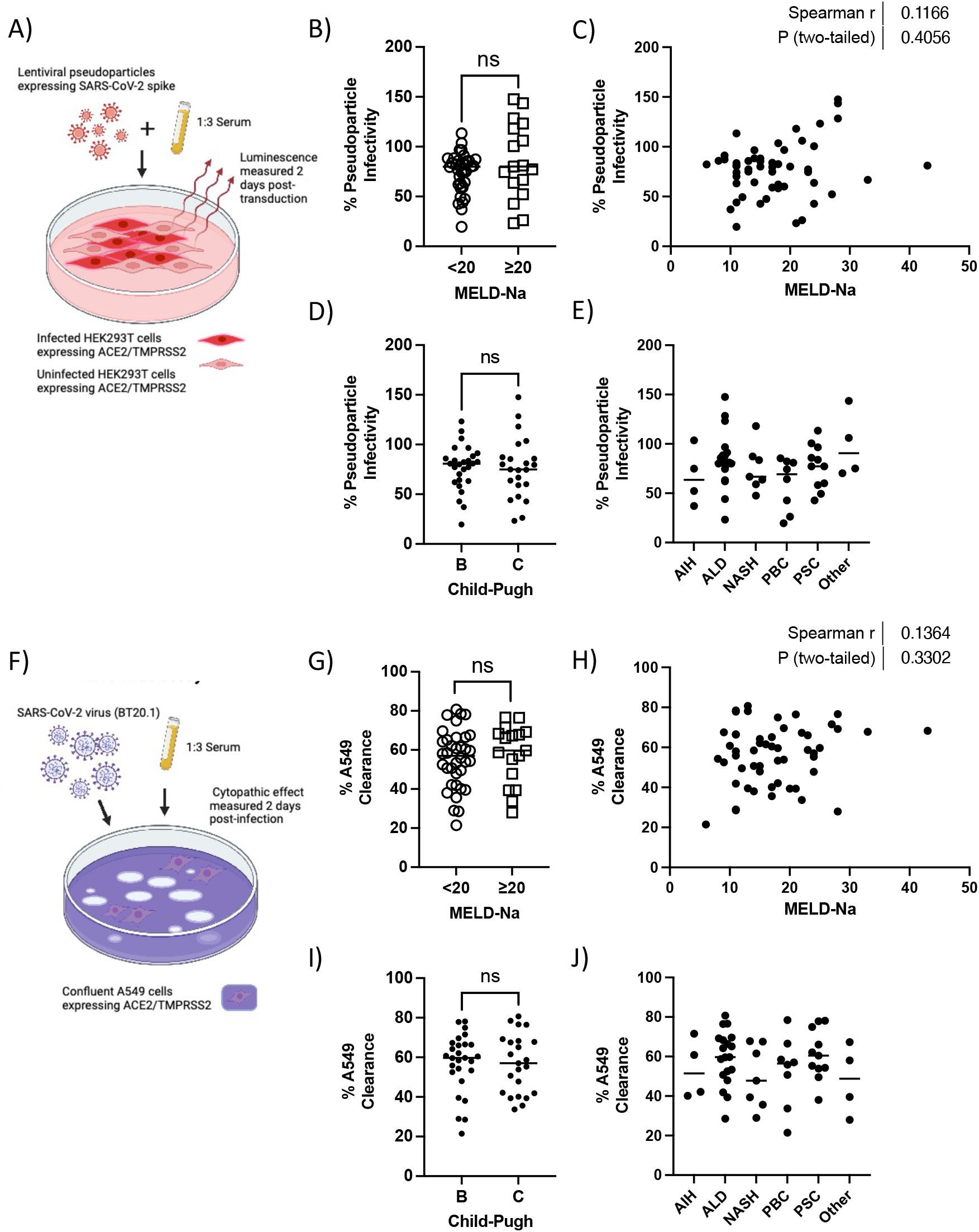
Serum from patients with varying severity of decompensated liver disease does not alter viral infectivity *in-vitro*. A) Schematic representation of the viral pseudoparticle (PP) assay. B) Infectivity of SARS-CoV-2 PPs in the presence of serum from patients with MELD-Na <20 versus MELD-Na ≥20. C) Spearman correlation analysis of MELD-Na score and SARS-CoV-2 PP infectivity. D) Infectivity of SARS-CoV-2 PPs in the presence of serum from patients grouped by Child-Pugh class. E) Comparison of infectivity of SARS-CoV-2 PPs in the presence of serum from patients grouped by aetiology. F) Schematic representation of the live virus assay using modified A549 lung epithelial cells. G) Infectivity of the SARS-CoV-2 BT20.1 strain (measured as clearance of modified A549 cells) in the presence of serum from patients with MELD-Na <20 versus MELD-Na ≥20. C) Spearman correlation analysis of MELD-Na score and infectivity of the SARS-CoV-2 BT20.1 strain. D) Infectivity of the SARS-CoV-2 BT20.1 strain in the presence of serum from patients grouped by Child-Pugh class. E) Comparison of infectivity of the SARS-CoV-2 BT20.1 strain in the presence of serum from patients grouped by aetiology. Data were analysed using the Mann-Whitney test. ns, not significant.

To confirm the findings from the pseudoparticle assay, a live virus SARS-CoV-2 (strain BT20.1) assay was established using modified A549 cells expressing ACE2 and TMPRSS2 (**Figure 1F**). Similar to the pseudoparticle assay, addition of diluted serum from patients with MELD-Na ≥20 did not result in significant differences in clearance of modified A549 cells compared to serum from patients with MELD-Na <20 (**Figure 1G**). MELD-Na <20 and MELD-Na ≥20 groups had a median percentage A549 clearance of 56.9% and 59.7% respectively, relative to the A549 clearance observed upon infection in the presence of DMEM alone (p=0.527). There was no significant correlation between clearance of modified A549 cells and MELD-Na scores across the full cohort (**Figure 1H**). Likewise, there was no significant difference in clearance of modified A549 cells between patients classified as either Child-Pugh class B or C, nor between patients with different disease aetiologies (**Figure 1I, J**).

No additional clinical parameters assessed, including age, gender, and frailty, were significantly associated with either the pseudoparticle infectivity or the clearance of modified A549 cells (**Supplementary Figure 2**). Despite the lack of association between viral infectivity and clinical parameters, it was evident that some individual serum samples were capable of influencing either pseudoparticle infectivity or clearance of modified A549 cells (**Figure 1B and G**). To explore factors contributing to this individual variation the serum proteome was compared between samples that inhibited and enhanced either pseudoparticle infectivity or A549 clearance. No significant differences were observed between samples that inhibited or enhanced pseudoparticle infectivity (data not shown). However, two proteins were significantly different between samples that inhibited or enhanced clearance of modified A549 cells (**Supplementary Figure 3A**). Alpha-2-macroglobulin (A2M) and sex hormone binding globulin (SHBG) levels were significantly associated with clearance of modified A549 cells across the full cohort (**Supplementary Figure 3B and C**) but they were not associated with pseudoparticle infectivity (**Supplementary Figure 3D and E**).

### Inflammatory, coagulation, and lipid metabolic pathways are dysregulated in the serum proteome of patients with more severe liver disease

Principal component analysis of the serum proteome (using the 171 proteins that were detected in >85% of samples) identified two main clinical parameters associated with serum proteome changes – aetiology and MELD-Na scores (**Figure 2A**). Principal component 1 separated samples on the basis of disease aetiology, indicating proteomic differences between patients with metabolic liver disease (ALD and NASH) versus autoimmune liver disease (PSC, PBC, AIH). Principal component 2 separated samples on the basis of MELD-Na scores, indicating that irrespective of disease aetiology a number of serum proteome changes occur in patients with high MELD-Na scores.

**Figure 2:**
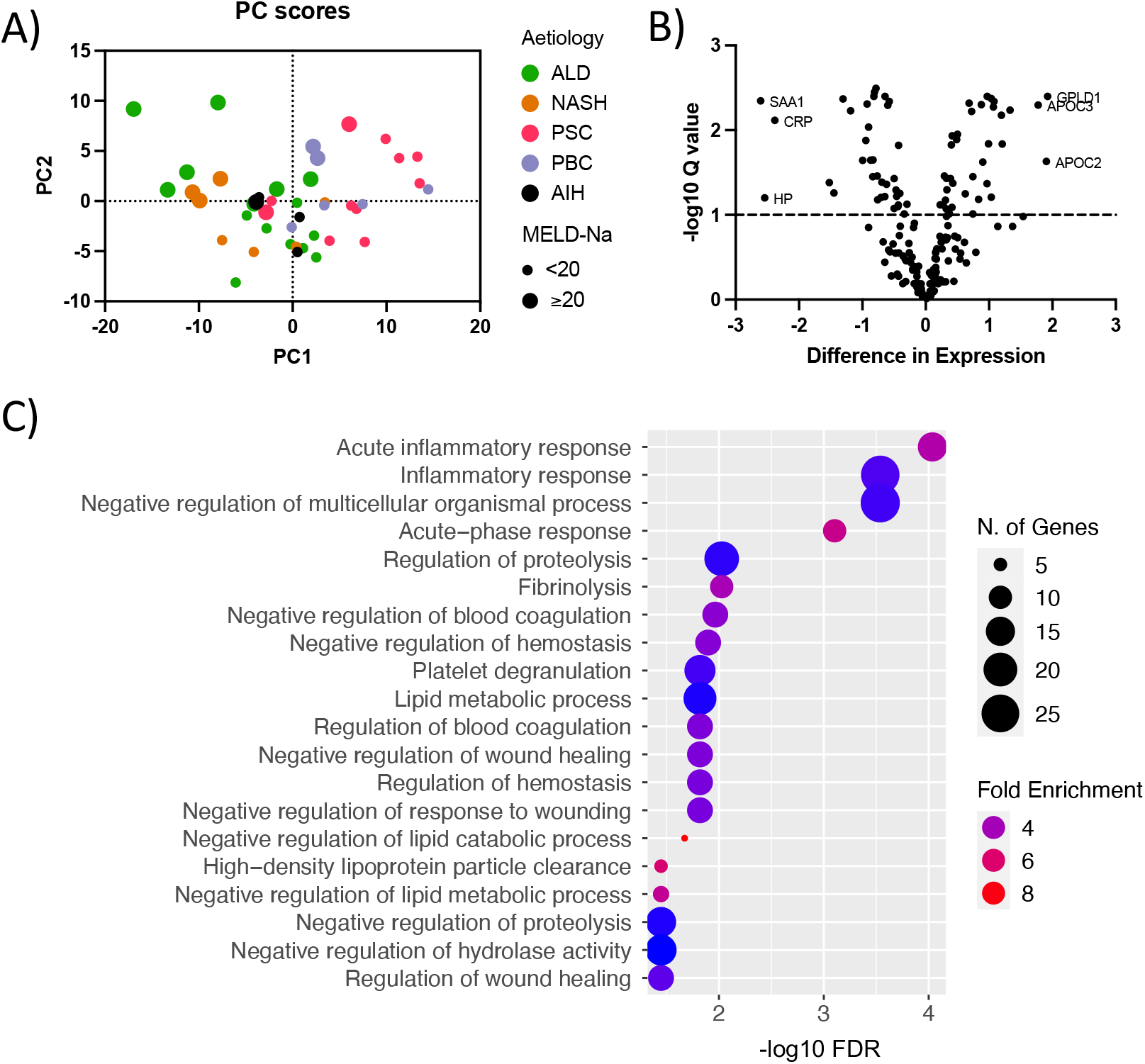
Dysregulated proteins in patients with more severe liver disease are involved in inflammatory, coagulation, and lipid metabolic pathways. A) Principal component analysis of mass spectrometry intensity values for 171 proteins passing quality filtering. Colours depict different disease aetiologies and dot size represents MELD-Na groupings. B) Volcano plot of the difference in expression and -log10 Q-value comparing ALD and NASH patients with MELD-Na <20 to those with MELD-Na ≥20 using unpaired t-test. C) Enriched GO biological processes amongst differentially expressed proteins from (B), classified as proteins with a Q-value <0.1. Terms are sorted by FDR with dot colour representing fold enrichment and dot size representing number of genes assigned to specific terms.

To explore the serum proteome changes associated with MELD-Na scores (and reduce the influence of disease aetiology), we restricted our analysis to patients with metabolic liver disease (ALD and NASH) and compared patients with MELD-Na <20 (n=12) and MELD-Na ≥20 (n=10, **Figure 2B**). A total of 72 proteins were differentially regulated between MELD-Na <20 and MELD-Na ≥20 patients (q-value < 0.1; **Figure 2B**). Amongst the proteins that increased the most in MELD-Na ≥20 patients there were a number associated with acute inflammatory responses, such as serum amyloid A1 (SAA1), C-reactive protein (CRP) and haptoglobin (HP). Amongst the proteins that decreased the most in MELD-Na ≥20 patients there were a number associated with lipid metabolism, such as apolipoprotein (APO)C2 and APOC3 and glycosylphosphatidylinositol specific phospholipase D1 (GPLD1). The 72 differentially regulated proteins were analysed using pathway analysis (**Figure 2C)**. Network analysis of the enriched GO Biological Process terms (FDR < 0.05) identified three main clusters of overlapping GO terms (**Figure 2C** and **Supplementary Figure 4**). These three clusters related to lipid metabolism, inflammatory responses and the regulation of coagulation and complement.

The majority of enriched GO terms were related to the regulation of coagulation and complement (**Figure 2C** and **Supplementary Figure 4**). Proteins upregulated in MELD-Na ≥20 patients from this cluster of GO terms included fibrinogen - and -chains (FGG, FGB), as well as two regulators of complement activation, glycoprotein Ib platelet subunit (GP1BA) and serpin family G member 1 (SERPING1) (**Supplementary Figure 4**). Proteins downregulated in MELD-Na ≥20 patients from this cluster of GO terms included coagulation factors II (F2) and XII (F12), kallikrein B1 (KLKB1), histidine rich glycoprotein (HRG), plasminogen (PLG), and carboxypeptidase B2 (CPB2) (**Supplementary Figure 4**). The reduction in F2, F12 and KLKB1 indicates a reduced ability to initiate blood coagulation via both the extrinsic and intrinsic pathways. At the same time, the reduction in PLG and increase in FGG and FGB indicates dysregulation of fibrinolysis processes. These proteomic changes mirror laboratory blood results where a trend toward increased platelets and a significant increase in INR was observed in from MELD-Na ≥20 patients (**Supplementary Figure 4E and F**).

The cluster of GO terms related to lipid metabolism included a large number of apolipoproteins (APOA2, APOC1, APOC2, APOC3, and APOD), as well as GPLD1, which were all lower in MELD-Na ≥20 patients (**Supplementary Figure 4**). This suppression of apolipoproteins is well documented in advanced liver disease and can predict survival of patients with liver failure [22, 23]. In addition to lipid metabolism, three of the four most significantly enriched GO terms were related to inflammatory processes (**Figure 2C**). Proteins upregulated in MELD-Na ≥20 patients from this cluster of GO terms included markers of endothelial cell activation (soluble adhesion molecules vascular cell adhesion molecule-1 (VCAM1) and intercellular adhesion molecule-1 (ICAM1)), markers of monocyte/macrophage activation (soluble CD163), and acute phase reactants (CRP, HP, orosomucoid (ORM)1, ORM2 and serpin family A member 3 (SERPINA3); **Supplementary Figure 4**). Proteins downregulated in MELD-Na ≥20 patients from this cluster of GO terms included coagulation factors (F2, F12), immunoregulatory serum glycoproteins (fibronectin 1 (FN1) and 2-HS Glycoprotein (ASHG)), and mannose-binding lectin 2 (MBL2). Despite the upregulation of proteins associated with acute inflammatory responses, no significant differences in circulating cytokine levels were evident (**Supplementary Figure 5**).

### Patients with more severe decompensated liver disease possess auto-antibodies capable of inhibiting type I IFN signalling

The results from the SARS-CoV-2 pseudoparticle and live virus experiments suggested there was no difference in intrinsic serum antiviral proteins in liver cirrhosis. However, the dysregulation of inflammatory pathways suggests that the ability of an individual’s immune cells to respond to viral infection may be altered. To explore whether the dysregulated inflammatory responses observed in the proteomic analysis were associated with type 1 IFN signalling in immune cells diluted serum (1/40 dilution) from decompensated patients with either MELD-Na <20 or MELD-Na ≥20 was pre-incubated with recombinant IFN-α2b (500 pg/ml), IFN-α8 (50 pg/ml) or IFN-β1a (100 pg/ml), and then assessed using the THP-1 dual reporter cells (**Figure 3**). Serum from patients with MELD-Na ≥20 had significantly reduced IFN-α2b and IFN-α8 activity in comparison to serum samples from patients with MELD-Na <20 (**Figure 3A and 3B**). The median IFN-α2b activity (relative to IFN-α2b alone) was 91.9% in the MELD-Na <20 group and 76.0% in the MELD-Na ≥20 group (p=0.002). The median activity for IFN-α8 in the MELD-Na <20 group versus the MELD-Na ≥20 group was 96.4% and 85.9%, respectively (p=0.038). There was no significant difference in IFN-β activity between the MELD-Na <20 group and the MELD-Na ≥20 group (median 112.9% versus 105.1%; **Figure 3C**).

**Figure 3:**
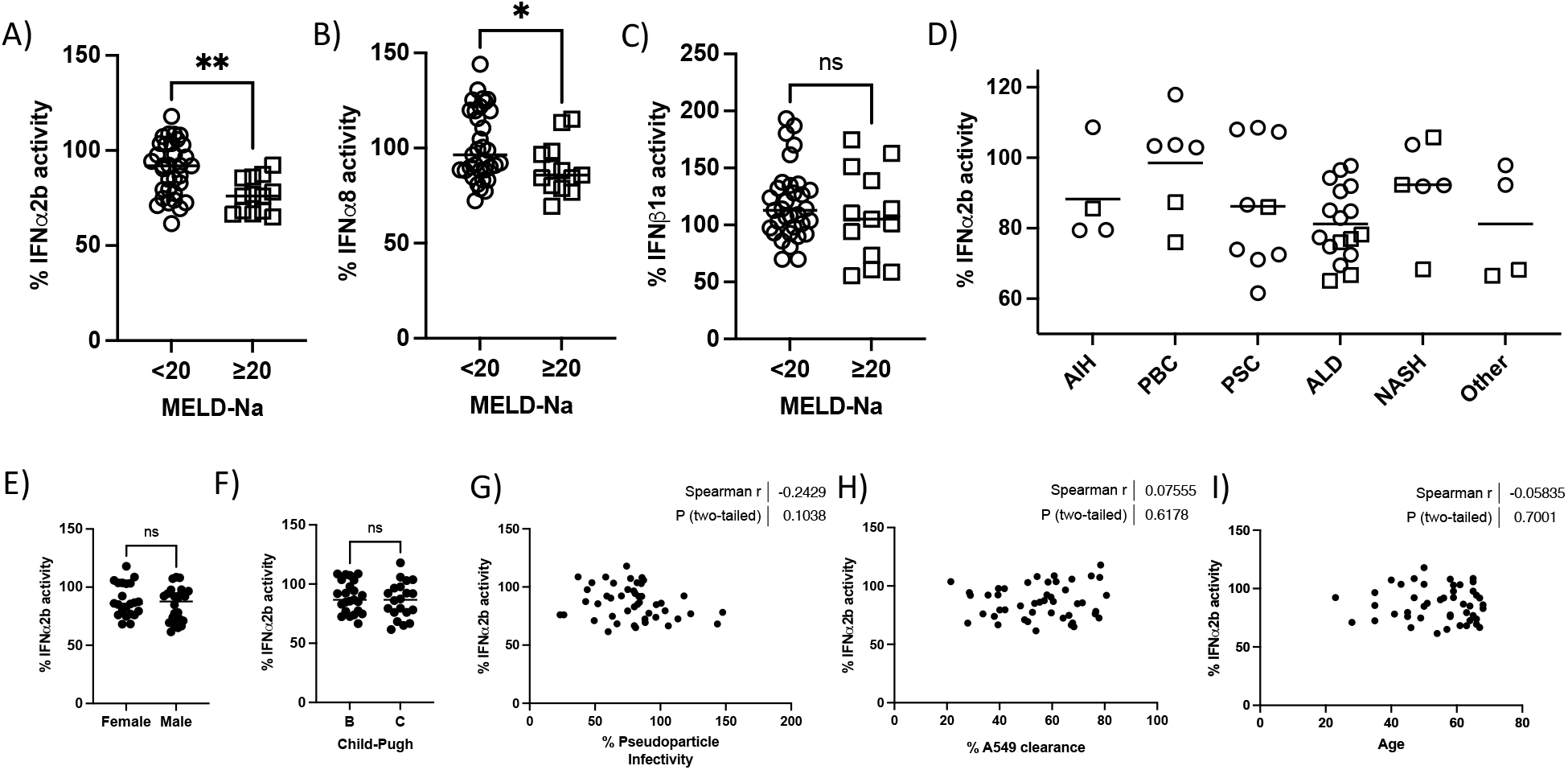
Patients with more severe decompensated liver disease display increased inhibition of IFNα signalling. A) Type I IFN activity following pre-incubation of IFNα2b (500 pg/ml) in the presence of a 1:40 dilution of serum from patients with MELD-Na <20 versus MELD-Na ≥20. B) Type I IFN activity following pre-incubation of IFN-α8 (50 pg/ml) in the presence of a 1:40 dilution of serum from patients with MELD-Na <20 versus MELD-Na ≥20. C) Type I IFN activity following pre-incubation of IFN-β1a (100 pg/ml) in the presence of a 1:40 dilution of serum from patients with MELD-Na <20 versus MELD-Na ≥20. D) Type I IFN activity following pre-incubation of IFNα2b (500 pg/ml) in the presence of a 1:40 dilution of serum grouped by disease aetiology. Circles designate individuals with MELD-Na <20 and squares designate individuals with MELD-Na ≥20. E) Type I IFN activity following pre-incubation of IFNα2b (500 pg/ml) in the presence of a 1:40 dilution of serum grouped by sex. F) Type I IFN activity following pre-incubation of IFNα2b (500 pg/ml) in the presence of a 1:40 dilution of serum grouped by Child-Pugh class. G-I) Spearman correlation analysis of type I IFN activity following pre-incubation of IFNα2b (500 pg/ml) in the presence of a 1:40 dilution of serum with viral psuedoparticle infectivity (G), live virus infectivity (H) and age (I). AIH, autoimmune hepatitis; ALD, alcoholic liver disease; NASH, non-alcoholic steatohepatitis; PBC, primary biliary cholangitis; PSC, primary sclerosing cholangitis. Data were analysed using the Mann-Whitney test. * *P*-value < 0.05; ** *P*-value < 0.01; ns = not significant.

This reduction in IFN-α2b activity observed in patients with MELD-Na ≥20 was not specific to a particular disease aetiology, although this conclusion is limited by the relatively small cohort size (**Figure 3D**). It is possible that disease aetiology may influence IFN-α2b activity, with the small number of PSC patients appearing to show a greater inhibition of type 1 IFN signalling even though most of these patients had MELD-Na scores <20 (**Figure 3D**). There was no significant variation in IFNα2b activity between sexes or patients classified as either Childs-Pugh class B or C (**Figure 3E and F**). There was also no correlation between IFNα2b activity and viral infectivity in either the pseudoparticle assay (**Figure 3G**) or the live virus assay (**Figure 3H**), likely due to the lack of cell types capable of producing IFNα in these model systems. There was also no correlation between IFNα2b activity and chronological age (**Figure 3I**).

As neutralising AAb against type I IFNs have been implicated in severe COVID-19 [9], we explored whether the inhibition of IFNα2b activity was mediated by auto-antibodies. Five serum samples that demonstrated inhibition of IFNα2b activity were selected for IgG purification, generating matched IgG-depleted serum and purified IgG. In comparison to matched whole serum the purified IgG showed significantly stronger inhibition of IFNα2b activity (mean IFNα2b activity with whole serum is 79.6% versus 52.6% with purified IgG; p=0.0001; **Figure 4A**). Removal of the IgG fraction restored IFNα2b activity, with the IgG-depleted serum showing a mean IFNα2b activity of 92.3% (**Figure 4A**).

**Figure 4:**
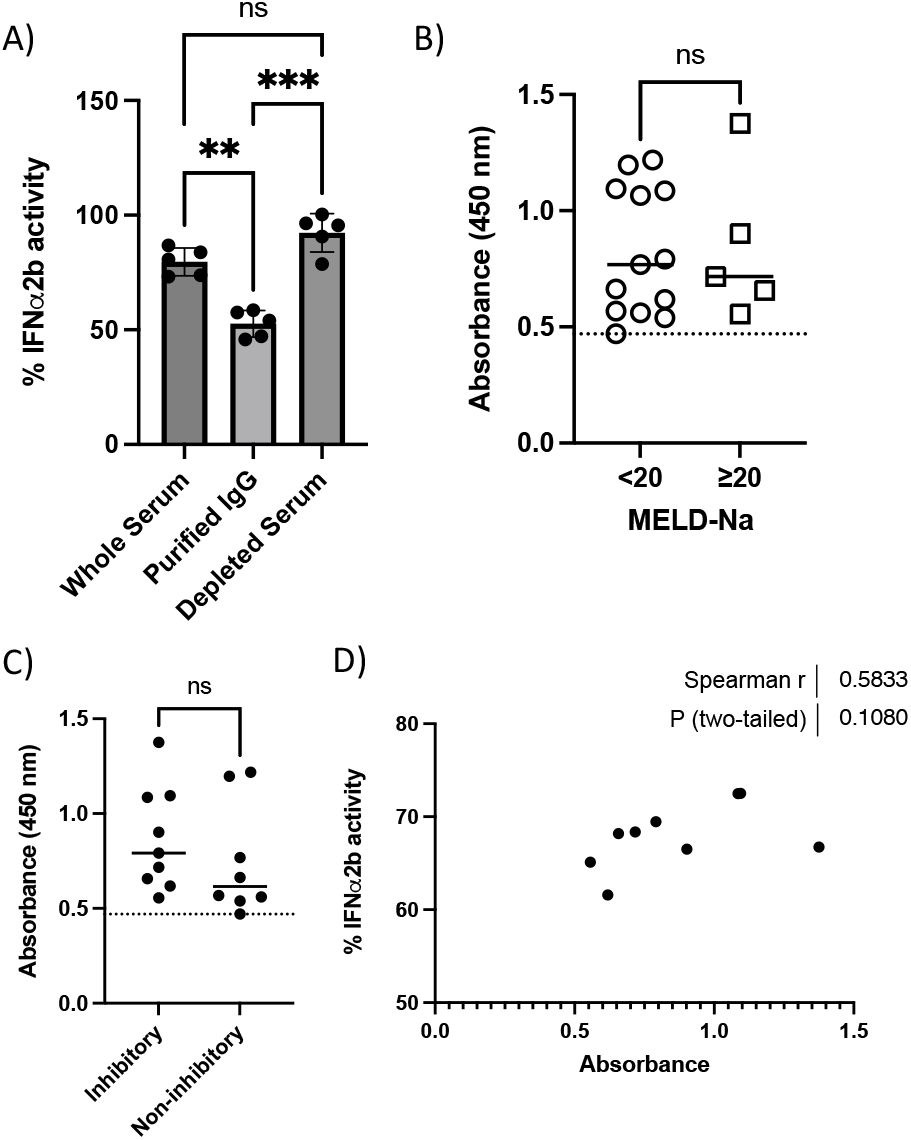
IgG auto-antibodies isolated from serum inhibit the activity of IFNα2b. A) Comparison of Type I IFN activity following pre-incubation of IFNα2b (500 pg/ml) in paired samples of whole serum, purified IgG and IgG depleted serum. B) Comparison of IFNα2b ELISA absorbances in serum samples from patients with MELD-Na <20 versus MELD-Na ≥20. Dotted line represents mean of healthy control serum. C) Comparison of IFNα2b ELISA absorbances in serum samples with evidence of inhibition of Type I IFN activity (‘inhibitory’) versus serum samples lacking inhibition of Type I IFN activity (‘non-inhibitory’). Dotted line represents mean of healthy control serum. D) Spearman correlation analysis of the type I IFN activity in ‘inhibitory’ samples following pre-incubation of IFNα2b (500 pg/ml) in the presence of a 1:40 dilution of serum and IFNα2b ELISA absorbance readings. Data were analysed using the repeated measures one-way ANOVA or unpaired t-test.**, *P*-value < 0.01; ***, *P*-value < 0.001; ns = not significant.

To determine whether the IgG was directly binding IFNα2b, as opposed to the type I IFN receptor, IFNα2b-binding was assessed using ELISAs. IgG reactivity against recombinant IFNα2b was evident in the majority of serum samples from decompensated patients, regardless of MELD-Na score (17/18 had absorbance readings greater than the mean of healthy control serum samples; **Figure 4B**). There was no significant difference in absorbance between patients with MELD-Na <20 and patients with MELD-Na ≥20 (**Figure 4B**). Samples were classified based on results from the IFN-α2b activity assays into either ‘inhibitory’ or ‘non-inhibitory’. Inhibitory samples were chosen from those with the greatest IFNα2b neutralising effect. The chosen samples had IFNα2b activity of <75% relative to the positive control. Non-inhibitory samples had a relative IFNα2b activity of >100%. While there was a slight increase in absorbance signal from the inhibitory samples, there was no significant difference in readings between inhibitory and non-inhibitory samples (**Figure 4C**), indicating that while detectable AAb against IFNα2b are present in most decompensated patients, they are not necessarily neutralising AAb. Furthermore, in the ‘inhibitory’ samples there was no significant correlation between purified IgG binding of IFNα2b and IFNα2b activity from the corresponding samples in the luciferase assay (**Figure 4D**). The lack of significant variation in ELISA absorbances between either the MELD-Na cohorts or ‘inhibitory’ versus ‘non-inhibitory’ samples, in combination with no significant correlation between IFNα2b activity and ELISA binding, demonstrates that both non-neutralising and neutralising AAb against type I IFN are present in patients with decompensated liver disease.

## Discussion

Pre-existing chronic liver disease is an important risk factor for severe COVID-19, especially in individuals with decompensated cirrhosis where the mortality rate in certain sub-groups is above 50% [8]. While vaccination is an important public health measure to reduce severe COVID-19, cirrhotic patients have poor responses to a variety of vaccines [24], and demonstrate reduced T cell responses and rapid loss of antibody responses to SARS-CoV-2 vaccines [25, 26]. Understanding the biological basis for susceptibility in this patient population is essential. In this study we examined intrinsic and innate antiviral responses to SARS-CoV-2 infection in unexposed decompensated cirrhotic patients, in order to understand alterations in viral immune responses in the absence of vaccination. Three individuals with strongly neutralising Ab responses against SARS-CoV-2 were identified and excluded from further analysis. Our results demonstrate that viral infectivity is not altered in the presence of serum from patients with decompensated liver cirrhosis, indicating that there is not a defect in intrinsic antiviral protein expression within the serum. In contrast, the serum proteome is significantly altered in patients with high MELD-Na scores and these patients possess neutralising AAb against type I interferon capable of inhibiting antiviral IFN signalling.

Neutralising AAb to type I IFNs have been identified as an important cause of severe COVID-19 in elderly individuals [9, 27]. We report that a 1/40 dilution of serum from patients with MELD-Na scores ≥20 significantly reduced the activity of 500 pg/mL IFNα2b and 50 pg/mL IFNα8 when compared to patients with less severe liver disease. Removal of IgG from these serum samples restored IFNα2b activity, confirming the presence of AAb with IFN neutralising activity. The level of neutralisation observed in our findings appears lower than that observed in the initial reports by Bastard and colleagues (reduction of IFN signalling to 15% or less of control) [9, 27]. The lack of standardised methodologies for detecting and quantifying neutralising type I IFN AAb is a limitation of our study. This limits the ability to directly compare the levels identified in decompensated cirrhotic patients to other studies of severe COVID-19 [28-31]. Our results are also limited by the fact that we detected the neutralising type I IFN AAb using a high sensitivity reporter cell line, and future research should validate the functional relevance of pre-existing type I IFN AAb on viral infectivity in this patient population. There is also a need to explore the relative frequency of type I IFN AAb in a wider range of chronic liver disease patients and age-matched health controls.

IFNα2b and IFNα8 neutralisation was more frequent in decompensated patients with high MELD-Na scores, there was no difference observed in IFNβ1a activity. IFNβ neutralising AAb have been rare or completely lacking in critical COVID patients possessing other neutralising type I IFN AAb [9, 32, 33]. IFNβ is a potent inducer of ISGs and the lack of neutralising AAb in this cohort could make it a potential treatment for COVID-19 in patients with decompensated liver disease. IFNβ has been used clinically in COVID-19 with positive results [33, 34]. The reason for the relative lack of AAb towards IFNβ is unknown but may stem from IFNβ being a more potent inducer of IFN stimulated genes (therefore it is expressed at lower concentrations). IFN neutralising AAb have been reported in a number of conditions with elevated IFN expression including autoimmune polyendocrinopathy syndrome [35], and system lupus erythematosus (SLE) [36, 37], as well as conditions such as myeloproliferative neoplasms where IFN therapeutics are used [38]. Repeated activation of type 1 IFNs has been reported in chronic liver diseases [39]. This chronic activation of type I IFNs may explain why neutralising AAb are present in cirrhotic patients.

Type I IFNs play an important role in the initial response of the host to a wide variety of viral infections, including respiratory tract infections [40, 41]. In SARS-CoV-2 infection rapid induction of IFN responses early in infection is required to limit SARS-CoV-2 replication [42-44]. Studies of the various IFNα subtypes have identified significant differences in the antiviral effects against SARS-CoV-2 [20]. We assessed neutralising AAb against IFNα2b and IFNα8, both of which show intermediate to high antiviral activity against SARS-CoV-2. This differential antiviral effect of different IFNα subtypes is thought to be due to differences in the induction of subsets of IFN stimulated genes in responding cells [20, 45]. Subtypes with high antiviral activity tend to induce robust expression of antiviral mediators such as *MX2, IFIT1, OAS2* and *RSAD2*. In contrast, subtypes with low antiviral activity preferentially induce gene signatures associated with cellular proliferation. The neutralisation of specific IFNα subtypes likely alters the dynamics between IFNα subtypes with high and low antiviral activity.

The presence of neutralising AAb against type I IFNs in patients with cirrhosis may act to reduce their initial response to infection and contribute to a more severe disease course. They are also of potential relevance for other viral infections. Bacterial infection is a risk factor for mortality in patients with liver cirrhosis [46]. However, data on the frequency of respiratory viral infections in this patient population is sparse. Respiratory viruses have been detected in approximately 20% of cirrhotic patients, often with bacterial co-infection, and is associated with poor clinical outcomes [47, 48]. The inhibition of IFN signalling may be a unrecognised component of the systemic immune dysregulation in chronic liver disease that could increase susceptibility to respiratory viral infection [49].

While serum from patients with high MELD-Na scores did not alter SARS-CoV-2 cell entry or viral replication in *in vitro* models, a high degree of inter-individual variation was observed in these assays. Serum contains a wide variety of proteins with both pro-viral and antiviral effects. We identified two serum proteins associated with reduced clearance of modified A549 cells upon infection with SARS-CoV-2. A2M is a highly abundant serum protein and has been identified as a possible inhibitor of SARS-CoV-2 entry, with an IC_50_ of 54.2μg/mL [50]. In human plasma, A2M has a concentration of approximately 1mg/mL, indicating it may play an important role in inhibiting SARS-CoV-2 entry. SHBG has been associated with mortality during SARS-CoV-2 infection in both men and women [51], although other studies have failed to identify this association with SHBG and clinical outcomes [52]. Our study did not include longitudinal follow-up of the patient cohort, and as such, we lacked data on subsequent infections with SARS-CoV-2. This limitation excluded the possibility of stratifying infection outcomes based on the antiviral activities evident in sera.

Patients with more severe end stage liver disease have dysregulation in inflammatory, coagulation and lipid metabolism pathways. Proteomic analysis of serum samples revealed a significant reduction in apolipoproteins and GPLD1 in patients with more severe liver disease. Dysfunctional lipid metabolism or dyslipidaemia has been documented in liver disease and is used as a predictor of disease severity [22, 23]. Dyslipidaemia has also been shown to have diagnostic and prognostic significance in certain bacterial and viral illnesses, including COVID-19 [53, 54]. A retrospective review from Li and colleagues revealed that HDL and APOA1 concentrations are significantly reduced in patients who had COVID-19 resulting in death [54]. A meta-analysis of observational studies suggested that both APOA1 and APOB could be protective against severe COVID-19 [55]. A large-scale multi-omics investigation of COVID-19 found that GPLD1 was significantly decreased in COVID-19 patients and correlated with more severe disease [56]. This pre-existing defect in lipid metabolism may predispose patients with liver cirrhosis to a more severe disease course upon SARS-CoV-2 infection.

The mass spectrometry data also revealed a significant dysregulation in proteins involved in the coagulation cascade in patients with high MELD-Na scores. This included a significant increase in the fibrinogen subunit chains and a significant reduction in proteins involved in the extrinsic and intrinsic coagulation pathways. Cirrhotic patients are known to be at risk of both thrombotic events as well as haemorrhagic events [57, 58]. Coagulopathies are also an important clinical manifestation of SARS-CoV-2 infection [59-61]. COVID-19 is characterised by prominent endothelial involvement in comparison to other viral pneumonias and thrombotic and micro-thrombotic events causing strokes, myocardial infarcts, and the more recently identified post-acute sequelae of COVID-19 or long COVID, are frequently observed [59-61].

Our data identifies possible vascular endothelial cell dysfunction and destruction associated with more severe liver disease, with significant upregulation of endothelial adhesion molecules, such as VCAM. Dysregulated systemic inflammation is an important driver of disease progression in liver cirrhosis [62]. While our results are limited to the serum proteome, previous research has documented alterations in immune function and responsiveness in peripheral blood immune cells in patients with liver cirrhosis [49].

Dysregulated inflammatory processes have been documented as a pathogenic mechanism in COVID-19. Conway and colleagues proposed a 3-step process of disease induction upon SARS-COV-2 infection; vascular endothelial cell dysfunction followed by hyper-inflammatory immune response and lastly excessive coagulation [59]. Collectively our results suggest the dysregulation of inflammatory, coagulation and lipid metabolism pathways, together with the presence of neutralising AAb targeting type I IFN may explain the increased likelihood of severe COVID-19 in chronic liver disease patients upon SARS-CoV-2 infection. Our findings are also of relevance to other viral infections in this patient population.

## Data Availability

All data produced in the present study are available upon reasonable request to the authors

## Acknowledgements

We wish to thank all participants for taking part in this research project and the support of the liver transplant team at St. Vincent’s University Hospital. This study was funded through the partnership between Science Foundation Ireland and the Department for the Economy (Northern Ireland) under the SFI COVID-19 Rapid Response Call (20/COV/8362). The cohort recruitment was funded by the Health Research Board (EIA-2017-013). The SARS-CoV-2 nucleocapsid ELISA was a kind gift from Prof. Sean Doyle, Maynooth University, developed from Science Foundation Ireland (SFI) COVID-19 Rapid Response Funding (20/COV/0048). The SARS-CoV2 anti-spike sero-assay work was funded by SFI Strategic Partnership Programme (20/SPP/3685). The SARS-CoV-2 pseudotype plasmids were kindly provided by Prof. Nigel Temperton, Medway School of Pharmacy, University of Kent. Human ACE2 plasmids were kindly provided by Dr Jacob Yount, Ohio State University. Modified A549 cells expressing human ACE2 and human TMPRSS2 were kindly provided by Dr Suzannah Rihn, MRC-University of Glasgow Centre for Virus Research.

## Ethics Statement

This study received ethics approval from St Vincent’s Hospital Group Ethics Board as per the 1975 Declaration of Helsinki guidelines.

---

**Supplementary Figure 1:**
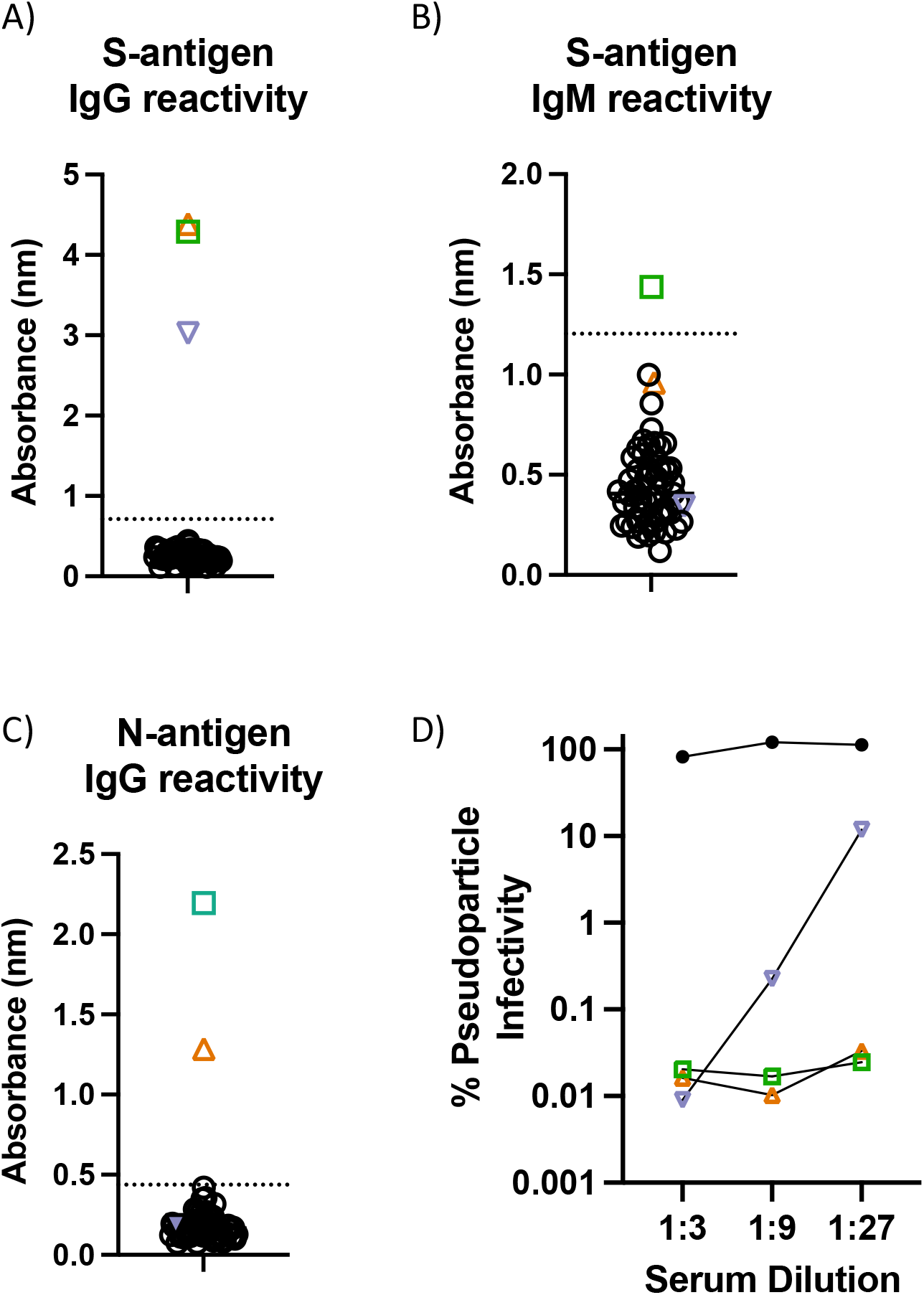
Serum reactivity against SARS-CoV-2 antigens indicative of previous exposure. A) S-antigen IgG reactivity, B) S-antigen IgM reactivity, and C) N-antigen IgG reactivity in the initial screening cohort of participants (n=60). The dotted line indicates the reference positive control sample absorbance. The three participants identified with serum reactivity against SARS-CoV-2 antigens are indicated by the coloured symbols (A-D). D) Assessment of the neutralisation ability of serum from the three participants with serum reactivity against SARS-CoV-2 antigens utilising the SARS-CoV-2 pseudoparticle assay at different serum dilutions. The black filled circles represents the neutralising ability of pooled unexposed healthy human serum.

**Supplementary Figure 2:**
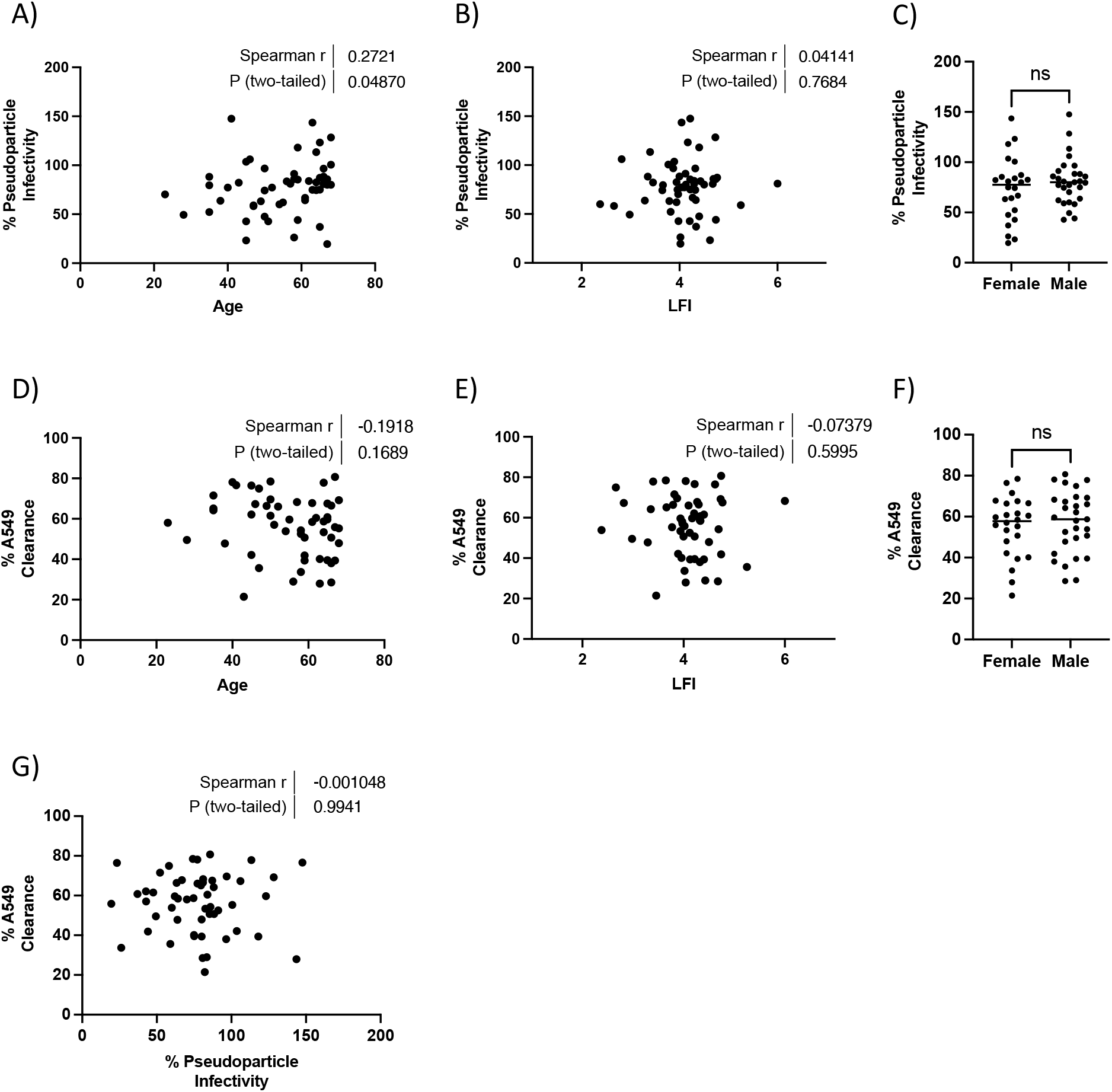
The impact of serum from patients with decompensated liver disease on viral infectivity *in-vitro* is not associated with clinically important parameters. A) Spearman correlation analysis of chronological age and infectivity of SARS-CoV-2 pseudoparticles (PP) in the presence of serum from patients with decompensated CLD. B) Spearman correlation analysis of liver frailty index (LFI) and infectivity of SARS-CoV-2 PP in the presence of serum from patients with decompensated CLD. C) Infectivity of SARS-CoV-2 PP in the presence of serum from patients grouped by sex. D) Spearman correlation analysis of chronological age and infectivity of the SARS-CoV-2 BT20.1 strain (measured as clearance of modified A549 cells) in the presence of serum from patients with decompensated CLD. E) Spearman correlation analysis of liver frailty index (LFI) and infectivity of the SARS-CoV-2 BT20.1 strain in the presence of serum from patients with decompensated CLD. F) Infectivity of the SARS-CoV-2 BT20.1 strain in the presence of serum from patients grouped by sex. G) Spearman correlation analysis of infectivity of SARS-CoV-2 PP versus infectivity of the SARS-CoV-2 BT20.1 strain in the presence of serum from patients with decompensated CLD. Data were analysed using the Mann-Whitney test. ns, not significant.

**Supplementary Figure 3:**
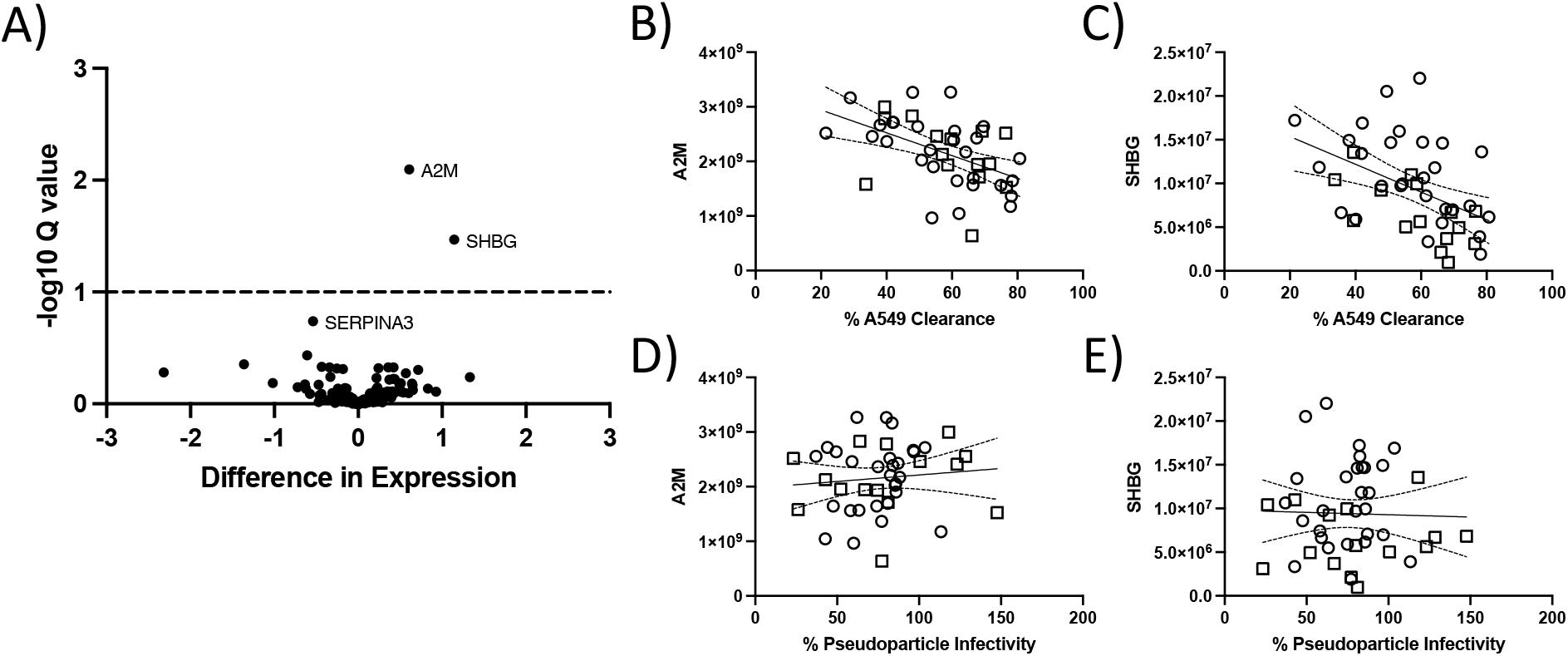
Association between the serum proteome from patients with decompensated liver disease and viral infectivity *in-vitro*. A) Volcano plot of the difference in expression and -log10 Q-value comparing individuals with low levels of modified A549 clearance upon infection with the SARS-CoV-2 BT20.1 strain (<50%) to those with high levels of A549 clearance (>65%) using unpaired t-test. B) Linear regression of infectivity of the SARS-CoV-2 BT20.1 strain in the presence of serum from patients with decompensated CLD plotted against alpha-2-macroglobulin (A2M) intensity values from mass spectrometry analysis. C) Linear regression of infectivity of the SARS-CoV-2 BT20.1 strain in the presence of serum from patients with decompensated CLD plotted against sex hormone binding globulin (SHBG) intensity values from mass spectrometry analysis. D) Linear regression of infectivity of SARS-CoV-2 pseudoparticles (PP) in the presence of serum from patients with decompensated CLD plotted against alpha-2-macroglobulin (A2M) intensity values from mass spectrometry analysis. E) Linear regression of infectivity of SARS-CoV-2 pseudoparticles (PP) in the presence of serum from patients with decompensated CLD plotted against sex hormone binding globulin (SHBG) intensity values from mass spectrometry analysis.

**Supplementary Figure 4:**
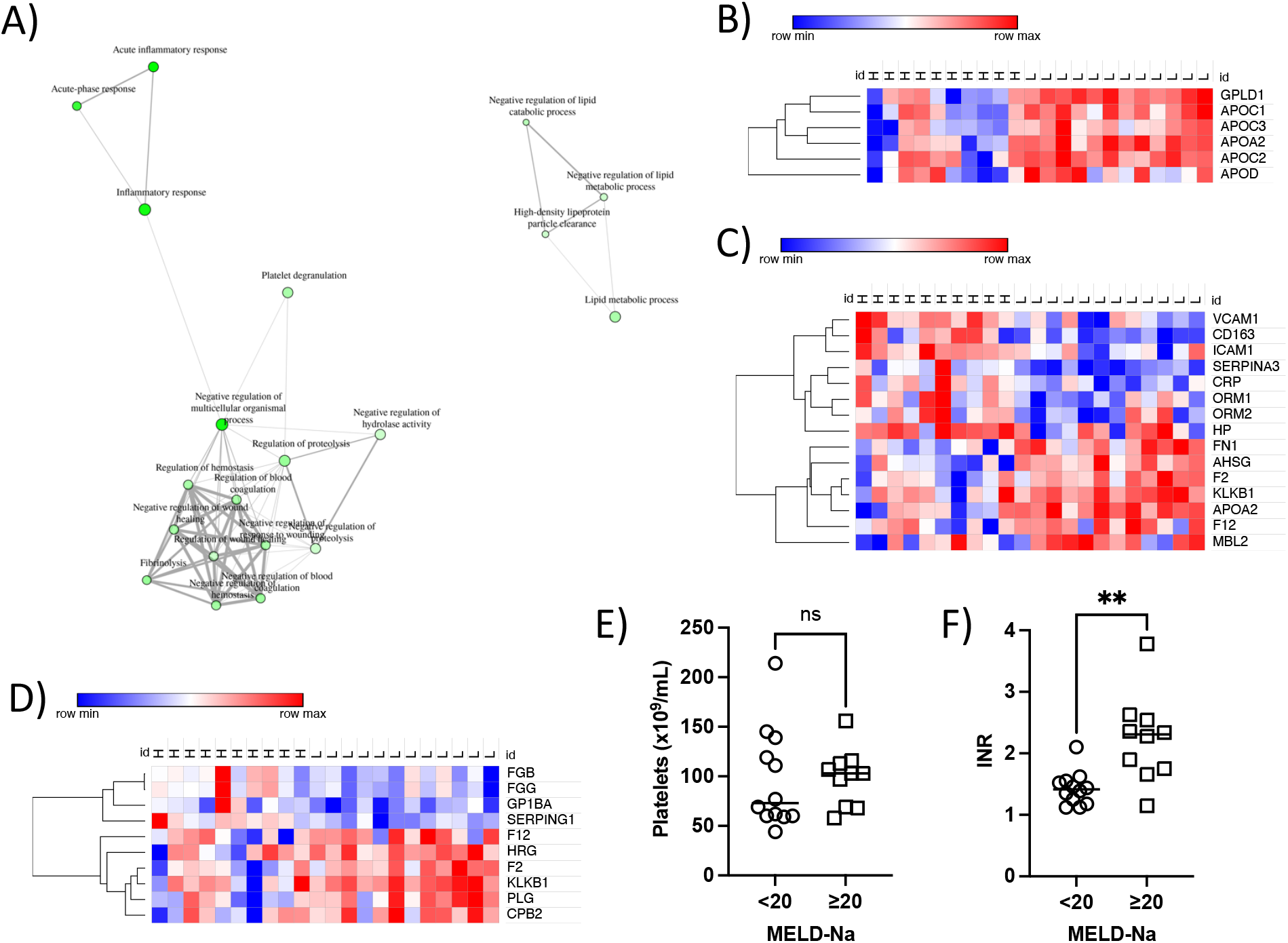
Network analysis identifies three core clusters of GO terms in patients with more severe liver disease. A) Network analysis of enriched GO terms amongst differentially expressed proteins (Q-value <0.1) comparing ALD and NASH patients with MELD-Na <20 to those with MELD-Na ≥20. Each node represents an enriched GO term, with related GO terms connected by a line, whose thickness reflects percent of overlapping genes. The size of the node corresponds to number of genes. B) Heatmap of proteins associated with the GO term ‘Negative regulation of lipid metabolic process’ in ALD and NASH patients with MELD-Na <20 (L) and MELD-Na ≥20 (H). C) Heatmap of proteins associated with the GO term ‘Acute inflammatory response’ in ALD and NASH patients with MELD-Na <20 (L) and MELD-Na ≥20 (H). D) Heatmap of proteins associated with the GO term ‘Fibrinolysis’ in ALD and NASH patients with MELD-Na <20 (L) and MELD-Na ≥20 (H). E) Platelets number comparing ALD and NASH patients with MELD-Na <20 to those with MELD-Na ≥20. F) INR comparing ALD and NASH patients with MELD-Na <20 to those with MELD-Na ≥20. Data were analysed using unpaired t-test. **, *P*-value < 0.01; ns, not significant.

**Supplementary Figure 5:**
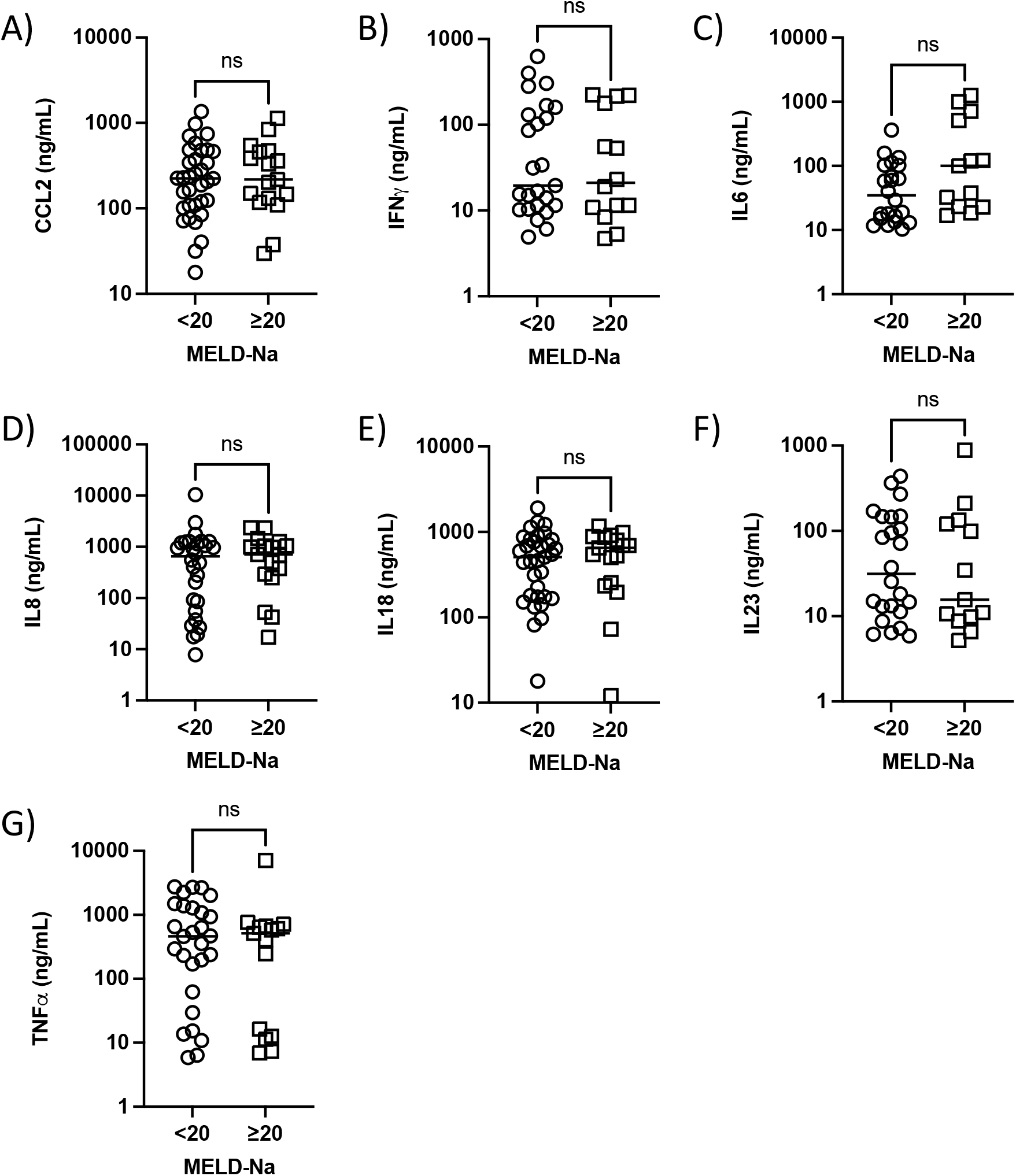
Circulating cytokine levels do not differ in patients with more severe liver disease. A-G) Serum cytokine levels in patients with MELD-Na <20 versus MELD-Na ≥20. A) CCL2, B) IFNγ, C) IL6, D) IL8, E) IL18, F) IL23, G) TNFα. Data were analysed using the Mann-Whitney test. ns, not significant.

## Notes

### Competing Interest Statement

The authors have declared no competing interest.

### Author Declarations

This study received ethics approval from St Vincents Hospital Group Ethics Board as per the 1975 Declaration of Helsinki guidelines.

